# Dynamics of neutralizing antibody titers in the months after SARS-CoV-2 infection

**DOI:** 10.1101/2020.08.06.20169367

**Authors:** Katharine H.D. Crawford, Adam S. Dingens, Rachel Eguia, Caitlin R. Wolf, Naomi Wilcox, Jennifer K. Logue, Kiel Shuey, Amanda M. Casto, Brooke Fiala, Samuel Wrenn, Deleah Pettie, Neil P. King, Helen Y. Chu, Jesse D. Bloom

## Abstract

Most individuals infected with SARS-CoV-2 develop neutralizing antibodies that target the viral spike protein. Here we quantify how levels of these antibodies change in the months following SARS-CoV-2 infection by examining longitudinal samples collected between ~30 and 152 days post-symptom onset from a prospective cohort of 34 recovered individuals with asymptomatic, mild, or moderate-severe disease. Neutralizing antibody titers declined an average of about four-fold from one to four months post-symptom onset. This decline in neutralizing antibody titers was accompanied by a decline in total antibodies capable of binding the viral spike or its receptor-binding domain. Importantly, our data are consistent with the expected early immune response to viral infection, where an initial peak in antibody levels is followed by a decline to a lower plateau. Additional studies of long-lived B-cells and antibody titers over longer time frames are necessary to determine the durability of immunity to SARS-CoV-2.

## Background

Within a few weeks of being infected with severe acute respiratory syndrome coronavirus 2 (SARS-CoV-2), individuals develop antibodies that bind to viral proteins [1-8]. A few weeks after symptom onset, sera from most infected individuals can bind to the viral spike and neutralize infection in vitro [5,7,9]. The reciprocal dilution of sera required to inhibit viral infection by 50% (NT_50_) is typically in the range of 100 to 200 at 3-4 weeks post-symptom onset [10], although neutralizing titers range from undetectable to >10,000 [2,5,9].

There are currently limited data on the dynamics of neutralizing antibodies in the months following recovery from SARS-CoV-2. For most acute viral infections, neutralizing antibodies rapidly rise after infection due to a burst of short-lived antibody-secreting cells, and then decline from this peak before reaching a stable plateau that can be maintained for years to decades by long-lived plasma and memory B cells [11,12]. These dynamics have been observed for many viruses, including influenza [13], RSV [14], MERS-CoV [15], SARS-CoV-1 [16,17], and the seasonal human coronavirus 229E [18].

Several recent studies have tracked antibody levels in individuals who have recovered from infection with SARS-CoV-2 for the first few months post-symptom onset [5,7,8,19-22]. Most of these studies have reported that over the first three months, antibodies targeting spike decline several fold from a peak reached a few weeks post-symptom onset [5,7,19], suggesting that the early dynamics of the antibody response to SARS-CoV-2 are similar to those for other acute viral infections.

Here we build on these studies by measuring both the neutralizing and binding antibody levels in serial plasma samples from 34 SARS-CoV-2-infected individuals across a range of disease severity with follow-up as long as 152 days post-symptom onset. On average, neutralizing titers decreased ~4-fold from ~30 to >90 days post-symptom onset. This decline in neutralizing titers was paralleled by a decrease in levels of antibodies that bind spike and its receptor-binding domain (RBD). Nonetheless, most recovered individuals still had substantial neutralizing titers at three to four months post-symptom onset.

## Methods

### Study population

Plasma samples were collected as part of a prospective longitudinal cohort study of individuals with SARS-CoV-2 infection. Individuals 18 years or older with laboratory confirmed SARS-CoV-2 infection were eligible for inclusion. Individuals who were HIV+ were excluded from this study due to concerns that antiretroviral treatment may affect our pseudotyped lentivirus neutralization assay. Individuals were recruited from three groups: inpatients, outpatients, and asymptomatics. Inpatients were hospitalized at Harborview Medical Center, University of Washington Medical Center, or Northwest Hospital in Seattle, WA and were enrolled while hospitalized. Outpatients were identified through a laboratory alert system, email and flyer advertising, and through identification of positive COVID-19 cases reported by the Seattle Flu Study [23]. Asymptomatic individuals in this study were recruited through outpatient testing and identified when they answered “None” on their symptom questionnaire. They were confirmed to be symptom-free for the first 30 days after diagnosis.

Participants or their legally authorized representatives completed electronic informed consent. Sociodemographic and clinical data were collected from electronic chart review and from participants via a data collection questionnaire (Project REDCap [24]) at the time of enrollment. The questionnaire collected data on the nature and duration of symptoms, medical comorbidities, and care-seeking behavior (**Supplementary Table 1**). Based on these data, individuals were classified by disease severity as *Asymptomatic, Symptomatic Non-Hospitalized*, and *Symptomatic Hospitalized*.

Individuals who were recruited as inpatients were enrolled during their hospital admission and had samples collected during their hospitalization. After hospital discharge, these participants subsequently returned to an outpatient clinical research site approximately 30 days after symptom onset for follow-up. In person follow-up only occurred if participants were asymptomatic as per CDC guidelines. Outpatients and asymptomatic individuals completed their enrollment, data collection questionnaire, and first blood draw at an outpatient visit approximately 30 days after symptom onset (or positive test for asymptomatic individuals). All participants subsequently were asked to return at day 60 and then at day 90 or 120 for follow-up.

The majority of samples collected from participants were from outpatient visits after recovery. However, the first sample from participant ID (PID) 13, the first three samples from PID 23, and the first six samples from PID 25 were collected during their hospitalizations.

For some of the analyses shown in Figures 1 and 2, samples from individuals were divided into three timepoints: ~30 days post-symptom onset (or post-positive test for asymptomatic individuals) (range: 22-48 days), ~60 days post-symptom onset or post-positive test (range: 55*-*79 days), and >90 days post-symptom onset or post-positive test (range: 94-152 days). Three individuals (PIDs: 2C, 23, and 25) had multiple samples in the first time range. For aggregated analyses that required samples be divided into these three groups, we only included the sample closest to 30 days post-symptom onset for those individuals. No individuals had multiple samples collected ~60 days post-symptom onset. One individual (PID: 12C) had two samples collected >90 days post-symptom onset. For this individual, we included only the latest sample collected in aggregated analyses that required classification into groups. The number of samples at each timepoint overall and for each disease severity classification is shown in **Table 1**. For analyses of fold-change, we required individuals to have a sample collected at the 30-day timepoint that had neutralizing antibody titers above our limit of detection (NT_50_ >20). The numbers of individuals included in the fold-change analyses are indicated in **Table 1**.

**Table 1:** Demographics of study participants and sample numbers for all disease severity

|  | **Asymptomatic**<br>**(N=7)** | **Symptomatic**<br>**Non-Hospitalized**<br>**(N=22)** | **Symptomatic**<br>**Hospitalized**<br>**(N=5)** | **Overall**<br>**(N=34)** |
| --- | --- | --- | --- | --- |
| **Age** |  |  |  |  |
| Median [Min, Max] | 62 [24, 79] | 43 [18, 76] | 54 [31, 64] | 45.5 [18, 79] |
| **Sex** |  |  |  |  |
| Male | 2 (28.6%) | 9 (40.9%) | 3 (60.0%) | 14 (41.2%) |
| Female | 5 (71.4%) | 13 (59.1%) | 2 (40.0%) | 20 (58.8%) |
| **Samples per Participant** |  |  |  |  |
| Median [Min, Max] | 2 [2-3] | 3 [2-3] | 3 [2-8] | 3 [2-8] |
| **Samples at each Timepoint**<br>**{Included in fold-change analyses}** |  |  |  |  |
| ~30 days | 5 {4} | 21 {20} | 4 {4} | 30 {28} |
| ~60 days | 7 {4} | 19 {17} | 3 {2} | 29 {23} |
| >90 days | 3 {1} | 15 {14} | 4 {3} | 22 {18} |
| **Days of Follow Up<sup>a</sup>** |  |  |  |  |
| Median [Min, Max] | 79 [60-131] | 104 [58-152] | 113 [76-121] | 103.5 [58-152] |

<sup>a</sup>Days between symptom onset date and collection date of last sample. For asymptomatic individuals, test date - calculated as Wednesday of the week of RT-qPCR test - was used in place of symptom onset date.

**Figure 1:**
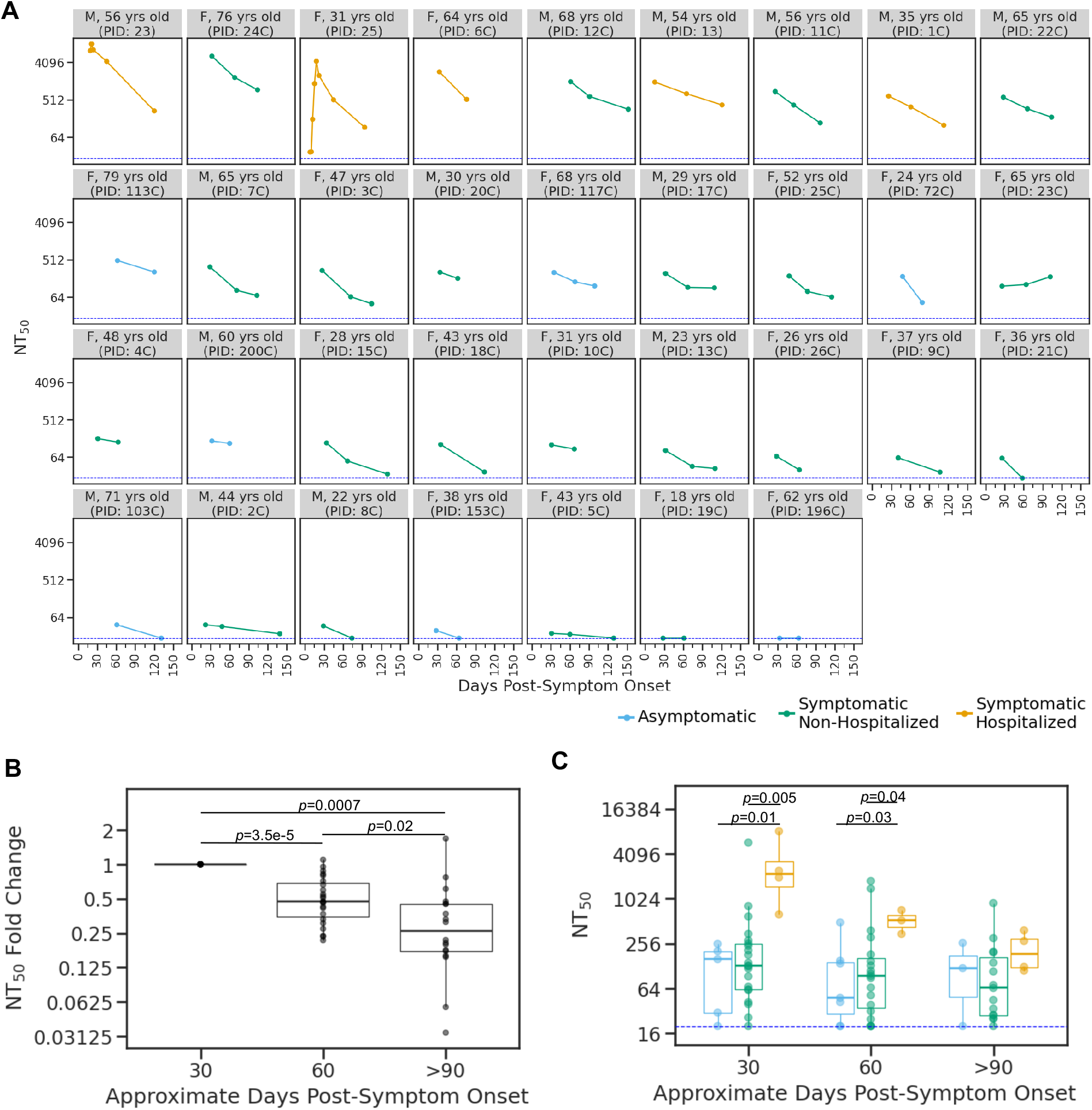
Change in neutralizing antibody titer over time. **A)** Neutralizing antibody titer at 50% inhibition (NT_50_) for each individual in the study, with facets colored according to disease severity (see key below plot). The facet titles indicate gender, age, and participant ID (PID). **B)** Fold change in NT_50_ compared to 30-day timepoint, including only individuals with a neutralizing sample at day 30. *P*-values were calculated using the Wilcoxon signed-rank test. **C)** Distribution of NT_50_s at the three timepoints, with boxplots colored by disease severity as in panel A. *P*-values are indicated when there is a significant difference (*p*≤0.05) between NT_50_s for different disease severity categories at a timepoint. *P*-values were calculated using the Wilcoxon rank-sum test.

**Figure 2:**
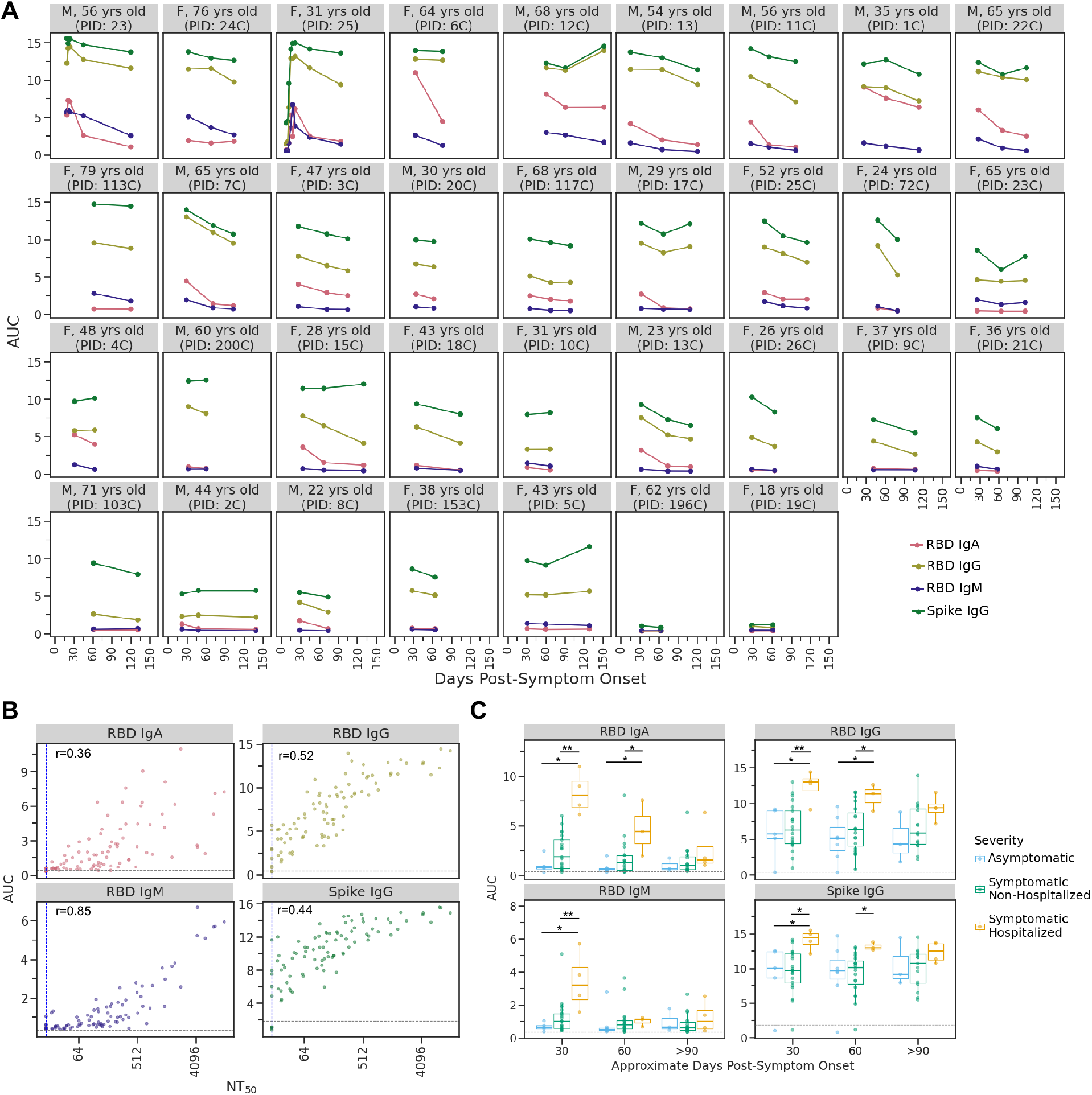
IgA, IgM, and IgG antibody binding titers overtime. **A)** Longitudinal binding antibody titers for each individual as quantified by area under the curve (AUC) of ELISA assays. Facets are arranged by maximal NT_50_ from top left to bottom right as in **Fig. 1A. B)** Correlation plots between AUC for each ELISA assay and neutralization titer (NT_50_) for all samples. The grey line in each facet indicates the AUC value for the negative control sample (2017-2018 sera pool) for each assay. **C)** For each antibody type measured, individuals who were symptomatic and required hospitalization as part of their care had significantly higher antibody levels during the first one to two months post-symptom onset. \**p*≤0.05, \*\**p*≤0.01. *P*-values were calculated using the Wilcoxon rank-sum test. As in **B**, the grey line in each facet indicates the AUC value for the negative control sample for each assay.

This study was approved by the University of Washington Human Subjects Institutional Review Board.

### Laboratory methods

Whole blood was collected in acid citrate dextrose tubes then spun down, aliquoted, and frozen at -20°C within 6 hours of collection. Prior to use in this study, plasma samples were heat inactivated at 56°C for 60 min and stored at 4°C. Some samples from the early timepoints were stored at -80°C after heat inactivation and underwent no more than two freeze/thaw cycles. Plasma samples were spun at 2000xg for 15 min at 4°C immediately prior to use to pellet platelets.

### Protein expression and purification

RBD and spike protein were produced in mammalian cells as previously described [24]. Briefly, the SARS-CoV-2 RBD and S-2P trimer [25] were produced in Expi293F cells grown in suspension using Expi293F expression medium (Life Technologies) at 33°C, 70% humidity, and 8% CO_2_, rotating at 150 rpm. The cultures were transfected using PEI-MAX (Polyscience) with cells grown to a density of 3.0 million cells per mL and cultivated for 3 days. Supernatants were clarified by centrifugation (5 minutes at 4000 rcf), addition of PDADMAC solution to a final concentration of 0.0375% (Sigma Aldrich, #409014), and a second centrifugation (5 minutes at 4000 rcf).

Proteins were purified from clarified supernatants via IMAC using Talon cobalt affinity resin (Takara), passing the flowthrough back over the column for a second binding step. The proteins were eluted in 20 mM Tris pH 8.0, 300 mM NaCl, and 300 mM imidazole and subsequently dialyzed into 50 mM Tris pH 7.0, 185 mM NaCl, 100 mM L-arginine, 4.5% (v/v) glycerol, and 0.75% (w/v) 3-[(3-cholamidopropyl)dimethylammonio]-2-hydroxy-1-propanesulfonate (CHAPS) (RBD) or 50 mM Tris pH 8.0, 150 mM NaCl, 0.25% L-histidine (spike trimer). SDS-PAGE was used to assess purity prior to flash freezing and storage at -80°C.

### ELISAs

IgG enzyme-linked immunosorbent assays (ELISAs) to spike and RBD were conducted as described previously [26], and were based on a published protocol that recently received emergency use authorization from New York State and the FDA [27,28]. Briefly, spike or RBD was diluted to 2 μg/mL in PBS, 50 μL of which was used to coat each well of 96 well Immunlon 2HB plates (Thermo Fisher; 3455) overnight at 4°C. The next day, plates were washed thrice with PBS containing 0.1% Tween 20 (PBS-T) using a plate washer (Tecan HydroFlex) and blocked for 1-2 hours at room temperature with 200 μL 3% non-fat dry milk in PBS-T. Plasma samples were diluted with five serial 3-fold dilutions in PBS-T containing 1% non-fat dry milk, starting at a 1:25 dilution. Each plate contained a negative control dilution series of pooled human sera collected from 2017-2018 (Gemini Biosciences, 100-110, lot H86W03J, pooled from 75 donors), and a CR3022 monoclonal antibody positive control dilution series starting at 1 ug/mL. Block was thrown off plates, and 100 μL diluted plasma was added to the plates and incubated at room temperature for 2 hours. Plates were then washed three times, and 50 μL of goat anti-human IgG-Fc horseradish peroxidase (HRP)-conjugated antibody (Bethyl Labs, A80-104P) diluted 1:3,000 in PBS-T containing 1% milk was added to each well and incubated for 1 hour at room temperature. Plates were again washed thrice with PBS-T, and 100 μL of TMB/E HRP substrate (Millipore Sigma; ES001) was then added to each well. After a 5 minute incubation, 100 μL 1N HCl was added, and the OD450 was read immediately on a Tecan infinite M1000Pro plate reader.

IgA and IgM RBD ELISAs were performed as described above, with the following changes. The IgA secondary antibody was Peroxidase AffiniPure Goat Anti-Human Serum IgA, α chain specific (Jackson Labs, 109-035-011), and the IgM secondary antibody was goat Anti-Human IgM (μ-chain specific)–Peroxidase antibody (Sigma Aldrich, A6907); both were diluted 1:3000 in PBS-T containing 1 % milk. For these ELISAs, plasma samples were run at six serial 4-fold dilutions starting at a 1:25 dilution, again with each plate containing a negative control dilution series (pooled human sera taken from 2017-2018).

AUC was calculated as the area under the titration curve after putting the serial dilutions on a log-scale.

### Neutralization assays

Neutralization assays were conducted using pseudotyped lentiviral particles as described in [29], with a few modifications. First, we used a spike with a cytoplasmic tail truncation that removes the last 21 amino acids (spike-Δ21). The map for this plasmid, HDM-SARS2-Spike-delta21, is in **Supplementary File 1** and the plasmid is available from Addgene (Plasmid #155130). We used a spike with a C-terminal deletion because, since publishing our original protocol [29], other groups have reported that deleting spike’s cytoplasmic tail improves titers of spike-pseudotyped viruses [30-33]. Indeed, we found that the C-terminal deletion increased the titers of our pseudotyped lentiviral particles without affecting neutralization sensitivity (**Supplementary Figure 1**).

For our neutralization assays, we seeded black-walled, clear bottom, poly-L-lysine coated 96-well plates (Greiner, 655936) with 1.25×10^4^ 293T-ACE2 (NR-52511) cells per well in 50 μL D10 media (DMEM with 10% heat-inactivated FBS, 2 mM l-glutamine, 100 U/mL penicillin, and 100 μg/mL streptomycin) at 37°C with 5% CO_2_. About 12 hours later, we diluted the plasma samples in D10 starting with a 1:20 dilution followed by 6 or 11 serial 3-fold dilutions (11 dilutions were used for samples from individuals we had previously measured as having high neutralizing antibody responses; PIDs: 13, 23, and 25). We then diluted the spike-Δ21 pseudotyped lentiviral particles 1 to 6 (1 mL of virus plus 5 mL of D10 per plate) and added a volume of virus equal to the volume of plasma dilution to each well of the plates containing the plasma dilutions. We incubated the virus and plasma plates for 1 hour at 37°C and then added 100 μL of the virus plus plasma dilutions to the cells ~16 hours after the cells were seeded.

At 50-52 hours post-infection, luciferase activity was measured using the Bright-Glo Luciferase Assay System (Promega, E2610) following the steps outlined in [29], except luciferase activity was measured directly in the assay plates and not transferred to opaque-bottom black plates. Two “no plasma” wells were included in each row of the neutralization plate and fraction infectivity was calculated by dividing the luciferase readings from the wells with plasma by the average of the “no plasma” wells in the same row. After calculating fraction infectivities, we used the `neutcurve` Python package (https://jbloomlab.github.io/neutcurve/) to calculate the plasma dilution that inhibited infection by 50% (IC50) by fitting a Hill curve with the bottom fixed at 0 and the top fixed at 1. NT_50_s for each plasma sample were calculated as the reciprocal of the IC50. Individuals whose plasma was not sufficiently neutralizing to interpolate an IC50 using the Hill curve fit were assigned an NT_50_ of 20 (the limit of our dilution series) for plotting in **Figures 1A, 1C**, and **2B** and for fold-change analyses in **Figure 1B**.

All samples were run in at least duplicate. We ran all samples from the same individual in the same batch of neutralization assays and on the same plate when possible. Each batch of samples included a negative control of pooled sera collected from 2017-2018 (Gemini Biosciences, 100-110, lot H86W03J, pooled from 75 donors), and one plasma sample known to be neutralizing (PID 4C, 30-day timepoint). These samples were used to confirm consistency between batches.

In an effort to help standardize comparisons between neutralization assays, we also ran our neutralization assay with a standard serum sample from NIBSC (Research Reagent for Anti-SARS-CoV-2 Ab, NIBSC code: 20/130). This sample had an NT_50_ of ~3050 (**Supplementary Figure 2**).

### Data availability

Raw data for each sample, including IC50, NT_50_, AUC and relevant demographic data (age, sex, disease severity, days post-symptom onset) are available as **Supplementary File 2**. Clinical data were analyzed in R version 3.6.0 (2019).

## Results

### Longitudinal plasma samples from a cohort of SARS-CoV-2 infected individuals

We enrolled 34 individuals following RT-qPCR-confirmed SARS-CoV-2 infection, of which five were symptomatic hospitalized, 22 were symptomatic non-hospitalized, and seven were asymptomatic (**Table 1, Supplementary Table 1**). This cohort included slightly more females than males (58.8% female overall) with ages ranging from 18 to 79. The age and sex distributions overall and based on disease severity are in **Table 1**. Five individuals had comorbidities. Information on participant race/ethnicity, symptoms, comorbidities, and level of medical care required is in **Supplementary Table 1**.

At least two samples were collected from all individuals in this study (median three samples) with the last sample collected between 58 and 152 days post-symptom onset (median 103 days). The majority (22/34) of individuals had their last sample collected >90 days post*-*symptom onset.

### Dynamics of neutralizing antibody titers over time

We used spike-pseudotyped lentiviral particles [29] to measure neutralizing antibody titers in the longitudinal plasma samples from all 34 individuals (**Figure 1A**). The vast majority of individuals (32/34) had detectable neutralizing antibody titers (NT_50_ >20) at roughly one month post*-*symptom onset, consistent with prior studies showing that most SARS-CoV-2 infected individuals develop neutralizing antibodies [5,7,9]. Qualitative inspection of **Figure 1A** shows that these titers modestly decreased for most individuals over the next few months, although the dynamics were highly heterogeneous across individuals.

To quantify the dynamics of neutralizing antibody titers over time, we calculated the fold change at ~60 and >90 days post-symptom onset relative to the ~30 day timepoint, excluding any individuals who lacked a 30-day sample or whose 30-day sample did not have detectable neutralizing titers (NT_50_ ≤20). Taken across all individuals, neutralizing titers significantly declined from 30 to 60 days, and again from 60 to 90 days (see legend of **Figure 1B** for details). At >90 days, the median neutralizing titer was reduced 3.8-fold relative to the 30 day value (**Figure 1B**). However, most individuals (27/34) still had detectable neutralizing titers at the last timepoint.

We compared the dynamics of neutralizing antibody titers between individuals with different disease severity (**Figure 1C**). Individuals with more severe disease tended to have higher neutralizing antibody titers during early convalescence, consistent with prior studies [5,34,35]. Specifically, at both ~30 and ~60 days post-symptom onset, individuals who required hospitalization had significantly higher neutralizing antibody titers than individuals who did not (**Figure 1C**). From ~30 to >90 days post-symptom onset, the NT_50_ for symptomatic hospitalized individuals decreased ~18-fold, which is significantly more than the ~3-fold decrease in the NT_50_ for non-hospitalized individuals (*p*=0.03, Wilcoxon rank-sum test) (**Supplementary Fig. 3**). By >90 days post-symptom onset, neutralization titers were not significantly different between disease severity groups (**Figure 1C**). At all timepoints, asymptomatic individuals had neutralization titers similar to those of symptomatic non-hospitalized individuals.

### Dynamics of spike- and RBD-binding antibodies over time

For all plasma samples, we also used ELISAs to measure IgA, IgM, and IgG binding to the RBD of spike, and IgG binding to the full spike ectodomain [27]. **Figure 2A** shows each individual’s IgA, IgM, and IgG binding antibody titers as quantified by area under the curve (AUC) of the ELISA readings (see Methods for detailed description). Like neutralizing antibody titers, these antibody binding titers tended to decrease over time, although there was substantial variation among individuals. All the ELISA-measured antibody-binding titers are clearly correlated with neutralizing antibody titers (**Figure 2B)**.

Individuals with severe disease had higher antibody binding titers at early timepoints. Specifically, individuals who were hospitalized as part of their care had higher IgG, IgA, and IgM binding responses than asymptomatic or symptomatic non-hospitalized individuals at ~30 days post-symptom onset (**Figure 2C**). By ~60 days post-symptom onset, anti-RBD IgM levels were no longer significantly different between severity groups, and by >90 days post-symptom onset, binding responses did not differ between severity groups for any antibody subtype. This trend is consistent with data in **Figure 1C** showing that neutralizing antibody responses were higher for individuals with more severe disease early during convalescence, but reached similar levels across all disease-severity groups by >90 days post symptom onset. Among all patients, regardless of disease severity, IgA and IgM levels decreased more than IgG levels from ~30 to >90 days post-symptom onset, consistent with other studies [7,8,19,22].

## Discussion

We have measured the dynamics of neutralizing antibody titers over the first three to four months following infection with SARS-CoV-2 in a well-characterized prospective longitudinal cohort of individuals across a range of disease severities. The titers of neutralizing antibodies declined modestly, with the titers at three to four months post-symptom onset generally about four-fold lower than those at one month. This decline in neutralizing antibodies was paralleled by a decline in antibodies binding to the viral spike and its RBD. This decline is generally similar in magnitude to that reported in several other recent studies of antibody dynamics in the months immediately following SARS-CoV-2 infection [5,7,19].

Individuals with more severe disease tended to have higher peak antibody responses at one to two months post-symptom onset, consistent with many other studies reporting higher early titers in severely ill SARS-CoV-2 infected individuals [5,6,34,35]. However, by three to four months post-symptom onset, neutralizing antibody titers among individuals with severe disease were no longer significantly higher than those of individuals with mild symptoms or even asymptomatic infections. Therefore, it seems possible that the large peak in antibody production in severely ill individuals wanes more dramatically than in milder cases, consistent with severe disease often leading to an exaggerated burst of short-lived antibody-secreting cells [36,37].

Importantly, most individuals in our study still had substantial neutralizing antibody titers at three to four months post-symptom onset. While some recent studies have interpreted a modest drop in titers in the first few months after infection as alarming, it is entirely consistent with antibody responses to other respiratory viruses. Acute infection is always associated with an initial peak in antibody titers due to a burst of short-lived antibody-secreting cells [38]. For many other infections, titers decline from this initial peak but then reach a stable plateau that is maintained for years or even decades by long-lived plasma cells and memory B cells that can be recalled during subsequent infections [12-16,18,39,40].

The limitations of this study include the small number of samples, particularly in the asymptomatic and symptomatic hospitalized groups, and recruitment of participants from a single study site, which potentially limits the generalizability of these results. Furthermore, since symptom-onset date relies on individual recollections, it is difficult to precisely match the timing of blood draws across all participants. Additionally, we only had follow-up to about four months post-symptom onset and only measured plasma antibody responses. Further studies over longer time frames and with direct interrogation of plasma and memory B cells will be necessary to determine longer term durability of immunity to SARS-CoV-2, as well as its relationship to protection against re-infection.

Despite these limitations, our study shows that titers of neutralizing and binding antibodies targeting SARS-CoV-2 spike remain detectable in most individuals out to >90 days post*-*symptom onset. While titers decline modestly from ~30 to >90 days post-symptom onset, we found that the dynamics of the antibody response to SARS-CoV-2 in the first several months following infection are consistent with what would be expected from knowledge of other acute viral infections [13-18].

## Data Availability

Raw data for each sample, including IC50, NT50, AUC and relevant demographic data (age, sex, disease severity, days post-symptom onset) are available as Supplementary File 2. Clinical data were analyzed in R version 3.6.0 (2019).

## Acknowledgements

We thank Marion Pepper for helpful input, Drs. David Koelle and Anna Wald for sharing reagents, and Andrea Loes for experimental assistance. We thank Ariana Magedson, Dylan McDonald and Nicholas Franko for assistance with enrollment. We additionally thank all our research participants in the HAARVI study for their generosity in participation.

## Funding

This research was supported by the following grants from the NIAID of the NIH: R01AI141707 (to J.D.B.) and F30AI149928 (to K.H.D.C.). Additional support was provided by the Bill & Melinda Gates Foundation (H.Y.C and OPP1156262 to N.P.K.). This study was also supported by an Investigators in the Pathogenesis of Infectious Disease Awards from the Burroughs Wellcome Fund (J.D.B.). J.D.B. is an Investigator of the Howard Hughes Medical Institute.

## Conflicts of Interest

NPK is a co-founder, shareholder, and chair of the scientific advisory board of Icosavax, Inc. HYC is a Consultant for Merck, Pfizer, and receives research funds from Cepheid, Ellume, Genentech, Sanofi-Pasteur. The other authors declare no conflicts of interest.

## Notes

### Author Declarations

This study was approved by the University of Washington Human Subjects Institutional Review Board.

